# Trends in excess cancer and cardiovascular deaths in Scotland during the COVID-19 pandemic 30 December – 20 April suggest underestimation of COVID-19 related deaths

**DOI:** 10.1101/2020.05.02.20086231

**Authors:** Jonine D. Figueroa, Paul Brennan, Evropi Theodoratou, Michael Poon, Karin Purshouse, Farhat Din, Kai Jin, Ines Mesa-Eguiagaray, Malcolm Dunlop, Peter Hall, David Cameron, Sarah Wild, Cathie Sudlow

## Abstract

Understanding the trends in causes of death for different diseases during the current COVID-19 pandemic is important to determine whether there are excess deaths beyond what is normally expected. Using the most recent report from National Records Scotland (NRS) on 29 April 2020, we examined the percentage difference in crude numbers of deaths in 2020 compared to the average for 2015-2019 by week of death within calendar year. To determine if trends were similar, suggesting underreporting/underdiagnosed COVID-19 related deaths, we also looked at the trends in % differences for cardiovascular disease deaths. From the first 17 weeks’ of data, we found a peak in excess deaths at week 14 of 2020, about four weeks after the first case in Scotland was detected on 1 March 2020-- but by week 17 these excesses had returned to normal levels, 4 weeks after lockdown in the UK began. Similar observations were seen for cardiovascular disease-related deaths. These observations suggest that the short-term increase in excess cancer and cardiovascular deaths might be associated with undetected/unconfirmed deaths related to COVID-19. Both of these conditions make patients more susceptible to infection and lack of widespread access to testing for COVID-19 are likely to have resulted in under-estimation of COVID-19 mortality. These data further suggest that the cumulative toll of COVID-19 on mortality is likely undercounted. More detailed analysis is needed to determine if these excesses were directly or indirectly related to COVID-19. Disease specific mortality will need constant monitoring for the foreseeable future as changes occur in increasing capacity and access to testing, reporting criteria, changes to health services and different measures are implemented to control the spread of the COVID-19. Multidisciplinary, multi-institutional, national and international collaborations for complementary and population specific data analysis is required to respond and mitigate adverse effects of the COVID-19 pandemic and to inform planning for future pandemics.

---

We read with great interest the <u>pre-print article by Lai et al^1^</u> which estimated that the direct and indirect effects of the COVID-19 emergency could lead to a 20% rise in cancer deaths in the UK and the US. Most people around the world are affected in some way by the COVID-19 pandemic, but the stresses are particularly heightened for people with, or being treated for, cancer. Recent data from the <u>ISARIC project in the UK^2^</u> showed that individuals with any malignancy are at increased risk of death following COVID-19 infection requiring hospital admission. In addition to being directly susceptible to COVID-19, there is growing concern that the lockdown in the UK, aimed at reducing virus transmission and allowing the NHS to cope with the number of patients with severe COVID-19 disease, may be having an unintentional adverse impact on patients with chronic illnesses, including cancer. The impact of reticence to attend primary care with symptoms of cancer, deferred referral from primary care, decreased capacity for optimal investigations to identify new cancers, and deferral of definitive cancer treatment, such as surgery, radiotherapy and chemotherapy, may all adversely impact on mode of presentation and cancer stage in the longer term. In the short term, management of late presentation of cancer with complications (e.g. bowel obstruction) is also associated with a higher morbidity and mortality. For instance, there is an approximately 10-fold greater 90 day-mortality for patients undergoing emergency colorectal cancer resections compared to those undergoing elective surgery (11.5% versus 1.7%)^3^. Furthermore, we have previously shown that elderly patients are the most likely to be disadvantaged by any measures constraining early diagnosis^4^.

Studying the trends in incidence and deaths for different diseases during the current COVID-19 pandemic is important to help policymakers, public health officials, clinicians and the wider public understand the intended and unintended effects of lockdown. Lai et al call for weekly data on cause-specific mortality, which has been noted to have delays in England, Wales and Northern Ireland, where in 2018 a review of cause of death showed increasing trends in the frequency of reporting delays for all causes, including neoplasms, with 23% of deaths due to neoplasms are often delayed beyond the 7-day reporting period^5^. By contrast, legislation in Scotland means that it has high-quality and timely registration of deaths, including cause of death, reported weekly^6^. Using the most recent report from National Records Scotland (NRS)^7^ on 29 April 2020, we examined the percentage difference in crude numbers of deaths in 2020 compared to the average for 2015-2019 by week of death within calendar year. We also examined the trends in reported cardiovascular disease deaths, another chronic disease for which there is great concern that there may be direct and indirect adverse effects of the COVID-19 emergency on mortality.

The left panel in the figure shows weekly cancer deaths registered in Scotland from 30 December 2019 to 20 April 2020 compared to the weekly average for the same week over the 5 previous years. During this time, we note a peak in excess deaths at week 14, about four weeks after the first case in Scotland was detected on 1 March 2020. The right panel shows a similar trend in the four-week moving average for both cancer and cardiovascular disease deaths after the first COVID-19 case was registered in Scotland at week 10, increasing to week 14, when the lockdown began, and declining thereafter. The observed short-term increase in cancer and cardiovascular deaths might be associated with undetected/unconfirmed deaths related to COVID-19, given the known issues of lack of widespread testing for infection and that both conditions increase susceptibility to the disease. These data are consistent with the increase in risk of death noted in the ISARIC UK report^2^, and reaffirm the importance of dynamic tracking of mortality data during the pandemic. The reduction in cardiovascular related deaths earlier in the year prior to week 10, compared with previous years, is likely due to a milder influenza season^7–8^. Whether trends in excess deaths stabilize beyond 20 April 2020 remains to be determined as new excesses might occur due to changes in health services and health seeking behaviors.

There are some important caveats to these data. The number of deaths where the cause has been reported as “other” deaths by NRS is currently higher for recent past weeks compared to the average for 2015-2019 by week of death within calendar year. Some deaths need an autopsy to determine their cause (for which results are available relatively quickly), while others need an inquest (which can take much longer). Some of “other” deaths reported by NRS will change and be redistributed to specific causes, some of which may be cancer deaths, although this is unlikely to change the general trend of excess cancer deaths. We examined crude numbers of deaths; more sophisticated age-adjusted and standardisation could provide more detailed insights. These data need ongoing monitoring for changes in the trends observed so far, especially if/when lockdown ends.

Incidence of cancer during the COVID-19 pandemic will undoubtedly be affected, especially in the short-term, with cancer screening services halted and a general hesitancy in seeking medical advice for problems perceived to be non-urgent. Whether this causes an excess in deaths will require long-term and detailed linkage studies. Cancer is a heterogeneous condition, with many different subtypes, different treatment regimens for molecularly distinct cancers at the same anatomical site (e.g. oestrogen receptor positive versus negative breast cancers), different referral patterns (e.g. breast, cervix and colorectal have screening programmes to detect cancer at earlier stages) and different survival patterns.

Understanding the impact of the COVID-19 pandemic, the changes to cancer services and cancer mortality require ongoing monitoring for the foreseeable future as different measures to address the COVID-19 pandemic are implemented. Multidisciplinary, multi-institutional, national and international collaborations for complementary and population specific data are needed to plan, respond and mitigate adverse effects to our populations.

Cancer diagnostic services are frequently over-stretched during normal times and ramping up both diagnostic and treatment services to cope with the backlog and concurrent new cases will require substantial resources. The current COVID-19 constraints on investigations and cancer treatment diagnosis will affect current incident cancers, but it will also affect patients yet to develop cancer, by creating future bottlenecks on stretched and imperfect diagnostic pathways. If the NHS is not adequately resourced, we could see new patients continue to be disadvantaged during the post-COVID catch up and plans to tackle this challenge are desperately needed. Continued monitoring of trends in incidence and mortality are required.

For now, it is important for cancer patients and all the public to know that while COVID-19 is a public health crisis, if they feel unwell or need advice the NHS is still open for normal business.

## Data Availability

Data are freely and publicly available

https://www.nrscotland.gov.uk/covid19stats

---

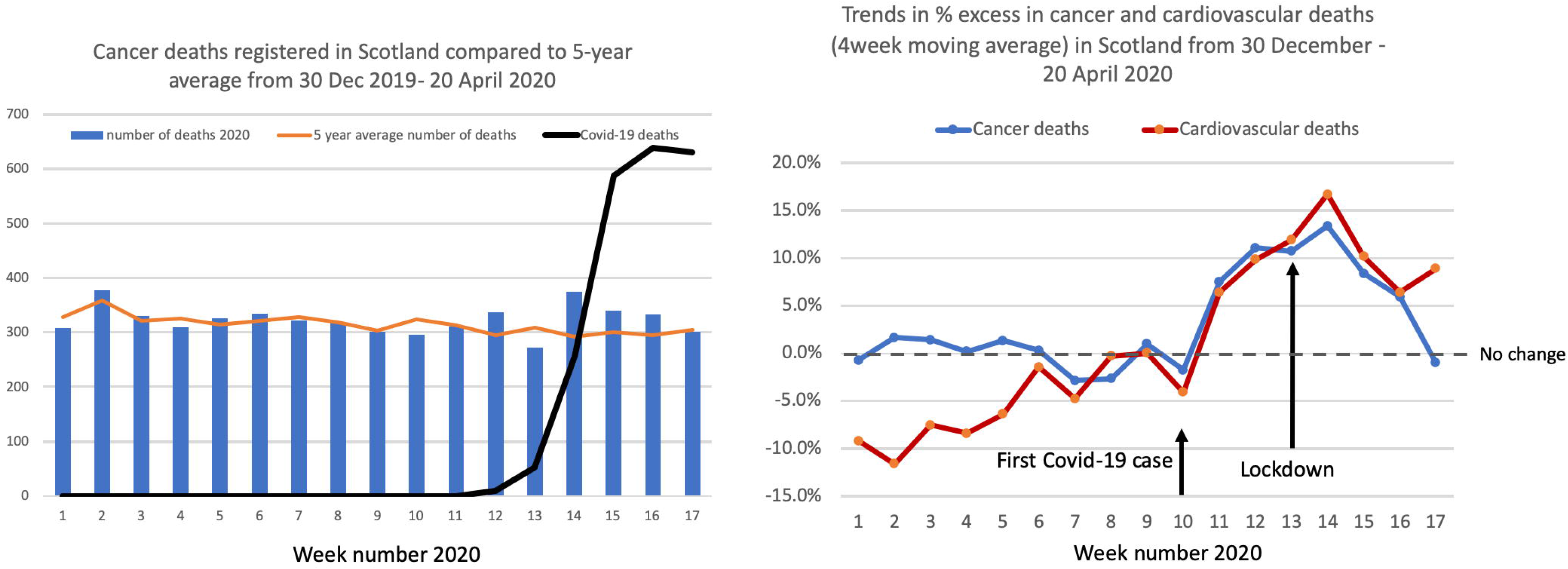
Data source figure 6 data fromNational Records Scotland download date 29 April 2020 https://www.nrscotland.gov.uk/covid19stats. Since the starts of 2020, a total of 23,281 deaths were reported in scotland. the X-axis on both figures are ther weeks of the year. in the left panel the Y-axis is the NRS reported number of deaths due to cancer compared to the 5-year average for similar time periods attributed to cancer (orange line). For reference, the black line shows the number of Covid-19 reported deaths. the right panel shows the 4-week moving average of excess % change in cancer (blue) and cardiovascular (red) deaths, with week 10 (1 march) the date when the first case of Covid-19 reported in scotland and the UK wide lockdown beginning week 13 (23 March). On average, “Other” deaths are about 23% of all deaths, but in recent weeks (from week 13) this is lower at 19% of all registered deaths. Over the last five years, cancer deaths have represented about 25% of all reported deaths.

